# Clinical utility of targeted SARS-CoV-2 serology testing to aid the diagnosis and management of suspected missed, late or post-COVID-19 infection syndromes: results from a pilot service

**DOI:** 10.1101/2020.07.10.20150540

**Authors:** Nicola Sweeney, Blair Merrick, Rui Pedro Galão, Suzanne Pickering, Alina Botros, Harry Wilson, Adrian W Signell, Gilberto Betancor, Mark Kia Ik Tan, John Ramble, Neophytos Kouphou, Sam Acors, Carl Graham, Jeffrey Seow, Eithne MacMahon, Stuart JD Neil, Michael H Malim, Katie Doores, Sam Douthwaite, Rahul Batra, Gaia Nebbia, Jonathan D Edgeworth

## Abstract

**OBJECTIVE:** Determine indications and clinical utility of SARS-CoV-2 serology testing in adults and children.

**DESIGN:** Prospective evaluation of initial three weeks of a daily Monday to Friday pilot SARS-CoV-2 serology service for patients.

**SETTING:** Early post “first-wave” SARS-CoV-2 transmission period at single centre London teaching hospital that provides care to the local community, as well as regional and national referral pathways for specialist services.

**PARTICIPANTS:** 110 (72 adults, 38 children, age range 0-83 years, 52.7% female (n=58)).

**INTERVENTIONS:** Patient serum from vetted referrals tested on CE marked and internally validated lateral flow immunoassay (LFIA) (SureScreen Diagnostics) detecting antibodies to SARS-CoV-2 spike proteins, with result and clinical interpretation provided to the direct care team.

**MAIN OUTCOME MEASURES:** Performance characteristics, source and nature of referrals, feasibility and clinical utility of the service, particularly the benefit for clinical decision-making.

**RESULTS:** The LFIA was deemed suitable for clinical advice and decision making following evaluation with 310 serum samples from SARS-CoV-2 PCR positive patients and 300 pre-pandemic samples, giving a sensitivity and specificity of 96.1% and 99.3% respectively. For the pilot, 115 referrals were received leading to 113 tests performed on 108 participants (sample not available for two participants); paediatrics (n=35), medicine (n=69), surgery (n=2) and general practice (n=2). 43.4% participants (n=49) had detectable antibodies to SARS-CoV-2. There were three main indications for serology; new acute presentations potentially triggered by recent COVID-19 infection e.g. PIMS-TS (n=26) and pulmonary embolism (n=5), potential missed diagnoses in context of a recent compatible illness (n=40), and making infection control and immunosuppression treatment decisions in persistently SARS-CoV-2 RNA PCR positive individuals (n=6).

**CONCLUSIONS:** This study shows acceptable performance characteristics, feasibility and clinical utility of a SARS-CoV-2 serology service using a rapid, inexpensive and portable assay for adults and children presenting with a range of clinical indications. Results correlated closely with a confirmatory in-house ELISA. The study showed the benefit of introducing a serology service where there is a reasonable pre-test probability, and the result can be linked with clinical advice or intervention. Experience thus far is that the volume of requests from hospital referral routes are manageable within existing clinical and laboratory services; however, the demand from community referrals has not yet been assessed. Given recent evidence for a rapid decline in antibodies, particularly following mild infection, there is likely a limited window of opportunity to realise the benefit of serology testing for individuals infected during the “first-wave” before they potentially fall below a measurable threshold. Rapidly expanding availability of serology services for NHS patients will also help understand the long-term implications of serostatus and prior infection in different patient groups, particularly before emergence of any “second-wave” outbreak or introduction of a vaccination programme.

**SUMMARY BOX:** *WHAT IS ALREADY KNOWN ON THIS TOPIC:* The mechanisms and utility of providing a SARS-CoV-2 (COVID-19) serology service is under evaluation. There are different technologies detecting antibodies against different SARS-CoV-2 proteins. Antibodies are known to appear from about 10 days after symptom onset but it is unclear how long they persist.

*WHAT THIS STUDY ADDS:* A SARS-CoV-2 serology service using a validated lateral flow immunoassay measuring antibodies against spike protein can be rapidly introduced with clinical benefit demonstrated for a broad range of individuals. Indications include ‘missed’ diagnoses where COVID-19 infection has been suspected but SARS-COV-2 RNA tests were either negative or not performed, conditions potentially triggered by COVID-19 such as pulmonary embolism, and predicting infectivity or immunity in patients with persistently detectable SARS-CoV-2 RNA. Testing is quick, simple to perform and inexpensive, however emerging evidence that antibodies fall rapidly particularly in mild disease, and the observed breadth of emerging indications highlight the urgent need for targeted testing with clinical interpretation provided on a case-by-case basis.

## INTRODUCTION

Infection with SARS-CoV-2 leads to production of a detectable antibody response in most people, however, the clinical utility of routine serological testing has been questioned.(1,2) There is uncertainty about what proportion of infected individuals produce serum antibodies, how long they persist for, and whether their detection provides protection against reinfection or disease manifestations upon re-exposure to the virus. These uncertainties, coupled with the fact that antibody testing for other respiratory viral infection is not standard practice and concerns regarding production and validation of rapidly developed new tests,(1,3) have led to hesitancy introducing them into widespread clinical practice.

From late May 2020 the UK government prioritised serological testing in NHS staff, reserving patient testing for those interested and undergoing other blood tests with a requirement for written consent. By that time our virology department had received many enquiries from different specialties asking whether SARS-CoV 2 infection might be contributing to patient presentation despite negative conventional RT-PCR testing.

We recently completed parallel validation of eight lateral flow immunoassay (LFIA) devices and two commercial ELISA platforms against an ELISA assay developed at King’s College London (KCL) that measures IgG, IgA and IgM against the main SARS-CoV-2 antigens (nucleocapsid (N) and spike (S) proteins and the S receptor binding domain (RBD). Viral neutralisation assays were also established alongside the in-house ELISA to correlate antibody titres with functional activity. Validation was initially performed on a cohort of patients presenting to Guy’s and St Thomas’ NHS Foundation Trust and showed that the accuracy of some of the lateral flow devices was comparable to our ELISA (paper submitted for publication).

We therefore submitted a formal request to the hospital Risk & Assurance Board sub-committee to provide a pilot clinical SARS-CoV-2 serology service for children and adults. The SureScreen LFIA was selected based on a range of factors including gaining confidence on performance and procurement during validation. Pilot approval was obtained on May 29^th^ 2020 following review of protocols and laboratory data including a further validation set reported here.

## METHODS

### SureScreen Diagnostics LFIA validation

Commercial LFIAs were selected for further validation based on results from previous head-to-head analyses.(4) Sensitivity and specificity experiments were designed with reference to MHRA and other validation guidance published at various times during the first wave, using serum samples from SARS-CoV-2 RNA positive (AusDiagnostics)(5) patients taken 14 or more (n=301) and 20 or more (n=204) days post onset of symptoms (POS) and 300 pre-pandemic samples. This included 200 stored serum samples and a panel of 100 stored acute and convalescent confounder samples taken from individuals with EBV, CMV, HIV and a range of other viral, bacterial and fungal pathogens. 95% confidence intervals were determined using the Wilson/Brown Binomial test. Sera from individuals diagnosed with seasonal coronaviruses were not available for testing. The research reagent for anti-SARS-CoV-2 Ab (NIBSC 20/130) obtained from the National Institute for Biological Standards and Control (NIBSC), UK, was used as a positive control for reproducibility and limit of detection experiments.

### Service delivery

Internal governance approval for service delivery was based on the laboratory validation data, clinical oversight, confirmation of an ability to request and report tests on electronic systems, a review of risks and their mitigation and agreement to report back on completion of the pilot. Service commenced on June 3rd and was delivered by scientists from the KCL Department of Infectious Diseases who had conducted all the LFIA validations. Tests were performed in and provided by the Guy’s and St Thomas’ Hospital Centre for Clinical Infection and Diagnostics Research (CIDR), located adjacent to hospital routine diagnostic virology and blood sciences laboratories on the St Thomas’ Hospital site.

Availability of SARS-CoV-2 serology service was communicated through clinical networks with requests vetted by the clinical virology team. Samples were requested as part of routine laboratory testing route and serology was performed once daily, Monday to Friday, using the SureScreen Diagnostics LFIA as per manufacturer’s instructions with two independent operators evaluating the result. A detectable band of either IgM or IgG (or both) was reported to the clinician as “antibodies detected”. Results were uploaded onto hospital electronic patient records as a scanned image of the lateral flow cassette with a written comment alongside telephoning where appropriate. Differential detection of IgM and IgG was not taken into account as part of verbal or written advice. Repeat testing was recommended when there was a high index of clinical suspicion and no antibodies were detected, or a weak positive IgM or IgG was the only observed band. A standard set of demographics, clinical information, request details and SARS-CoV-2 PCR results were recorded for each participant and stored in a clinical database. Clinicians using the service were contacted for informal feedback and their views on utility. Sera were batched for testing on the KCL ELISA platform with additional assessment for neutralizing antibodies where appropriate. This was undertaken as a further level of validation, but at a time remote from clinical decision making.

### ELISA

High-binding ELISA plates (Corning, 3690) were coated with antigen (N, S) at 3 µg/mL (25 µL per well) in PBS. Wells were washed with PBS-T (PBS with 0.05% Tween-20) and then blocked with 100 µL 5% milk in PBS-T for 1 hr at room temperature. Wells were emptied and sera diluted at 1:50 in milk was added and incubated for 2 hr at room temperature. Control reagents included CR3009 (2 µg/mL), CR3022 (0.2 µg/mL), negative control plasma (1:25 dilution), positive control plasma (1:50) and blank wells. Wells were washed with PBS-T. Secondary antibody was added and incubated for 1 hr at room temperature. IgM was detected using goat-anti-human-IgM-HRP (1:1,000) (Sigma: A6907), IgG was detected using goat-anti-human-Fc-AP (1:1,000) (Jackson: 109-055-043-JIR). Wells were washed with PBS-T and either Alkaline Phosphatase (AP) substrate (Sigma) was added and read at 405 nm (AP) or 1-step TMB substrate (Thermo Scientific) was added and quenched with 0.5 M H_2_SO_4_ before reading at 450 nm (HRP). Antibodies were considered detected if OD values were 4-fold or greater above background.

### Neutralising antibody assay

Serial dilutions of serum samples were prepared with DMEM media and incubated with pseudotyped HIV virus incorporating the SARS-Cov2 spike protein(6) for 1-hour at 37°C in 96-well plates. Next, HeLa cells stably expressing the ACE2 receptor (provided by Dr James Voss, The Scripps Research Institute) were added and the plates were left for 72 hours. Infection level was assessed in lysed cells with the Bright-Glo luciferase kit (Promega), using a Victor™ X3 multilabel reader (Perkin Elmer). The ID50 for each sera was calculated using GraphPad Prism.The ID_50_ for each sera was calculated using GraphPad Prism. Neutralisation titres were classified as low (50-200), moderate (201-500), high (501-2000), or potent (2001+).

### Patient and public involvement

Patients were not involved in the development of the study or its outcome measures, conduct of the research, or preparation of the manuscript.

## RESULTS

Comprehensive LFIA validation was performed using serum samples from 301 PCR-confirmed SARS-CoV-2 positive individuals collected 14 or more days POS and 300 pre-pandemic serum samples including 100 (acute and convalescent) from patients with a range of other infections that could give a false positive result (table 1a). 168 (of the 301) samples were specifically evaluated head-to-head with an in-house ELISA for IgM and IgG to N, S and RBD (supplementary figure 1). Sensitivity at 14 and 20 days or more POS was 94.4% and 96.1% respectively and specificity was 99.3% (table 1b). Limit of detection based on visual inspection of LFIA bands by two operators was determined using the NIBSC reference standard to a dilution of 1 in 500, consistent with the expected limit of detection of the NIBSC in-house assay(7).

**Table 1a:** Specificity of SureScreen lateral flow immunoassay

| Panel | Samples tested (n) | Positive (n) | Specificity (95% CI) |
| --- | --- | --- | --- |
| Pre-pandemic (from March 2019) | 200 | 1* | 99.5 % |
| Cytomegalovirus (CMV) | 8 | 0 | 100 % |
| Epstein-Barr virus (EBV) | 10 | 0 | 100 % |
| Hepatitis A virus | 8 | 0 | 100 % |
| Hepatitis B virus | 7 | 0 | 100 % |
| Hepatitis C virus | 5 | 0 | 100 % |
| Human immunodeficiency virus (HIV) | 9 | 0 | 100 % |
| Kaposi's sarcoma herpesvirus 1/2 | 5 | 0 | 100 % |
| Measles virus | 6 | 1* | 83.3 % |
| Mumps | 9 | 0 | 100 % |
| Mycobacterium | 1 | 0 | 100 % |
| Parvovirus | 7 | 0 | 100 % |
| Pneumocystis pneumonia | 4 | 0 | 100 % |
| Rubella virus | 5 | 0 | 100 % |
| Syphilis virus | 4 | 0 | 100 % |
| Toxoplasma gondii | 7 | 0 | 100 % |
| Varicella zoster virus | 5 | 0 | 100 % |
| Confounder samples (all) | 100 | 1 | 99.0 % |
| Overall | 300 | 2 | 99.3% (97.6-99.8) |

\*False positives were single (either IgM or IgG) weak bands. A similar result in the pilot triggered a request for a repeat sample to test.

**Table 1b:** Sensitivity of SureScreen lateral flow immunoassay

| Panel | Samples tested (n) | Positive (n) | Sensitivity (95% CI) |
| --- | --- | --- | --- |
| SARS-CoV-2-positive 14+ days POS | 301 | 284 | 94.4 % (91.1-96.4) |
| SARS-CoV-2-positive 20+ days POS | 204 | 196 | 96.1 % (92.4-98.0) |

POS= post onset of symptoms

Overall 49/108 (45%) participants had detectable IgG and/or IgM SARS-CoV-2 antibodies on their first serum sample that was communicated to clinicians as “antibodies detected” (table 2). 38/49 (78%) had IgM and 48/49 (98%) had IgG bands. Five participants with a high index of suspicion but no detectable antibodies had a further serum sample tested at least one week after initial testing. All repeat samples had no detectable antibodies. Rationale for testing broadly fell into three referral categories. First, acute presentations with new symptoms potentially triggered by SARS-CoV-2 infection. This included suspected cases of Paediatric Inflammatory Multisystem Syndrome Temporally associated with SARS-CoV-2 (PIMS-TS) (n=29), plus adults (n=27) and children (n=6) presenting with other clinical syndromes including thrombotic events such as strokes and pulmonary emboli (collectively called COVID-19 syndromes). Second, suspected “missed” diagnoses in individuals with a (recent) COVID-19 compatible illness who either never had an RNA test performed (n=18) or viral RNA was not detected in respiratory specimens (n=22). Third, those for whom antibody detection made a significant contribution to decisions on infection control management or immunosuppressive treatment (n=6).

**Table 2:**
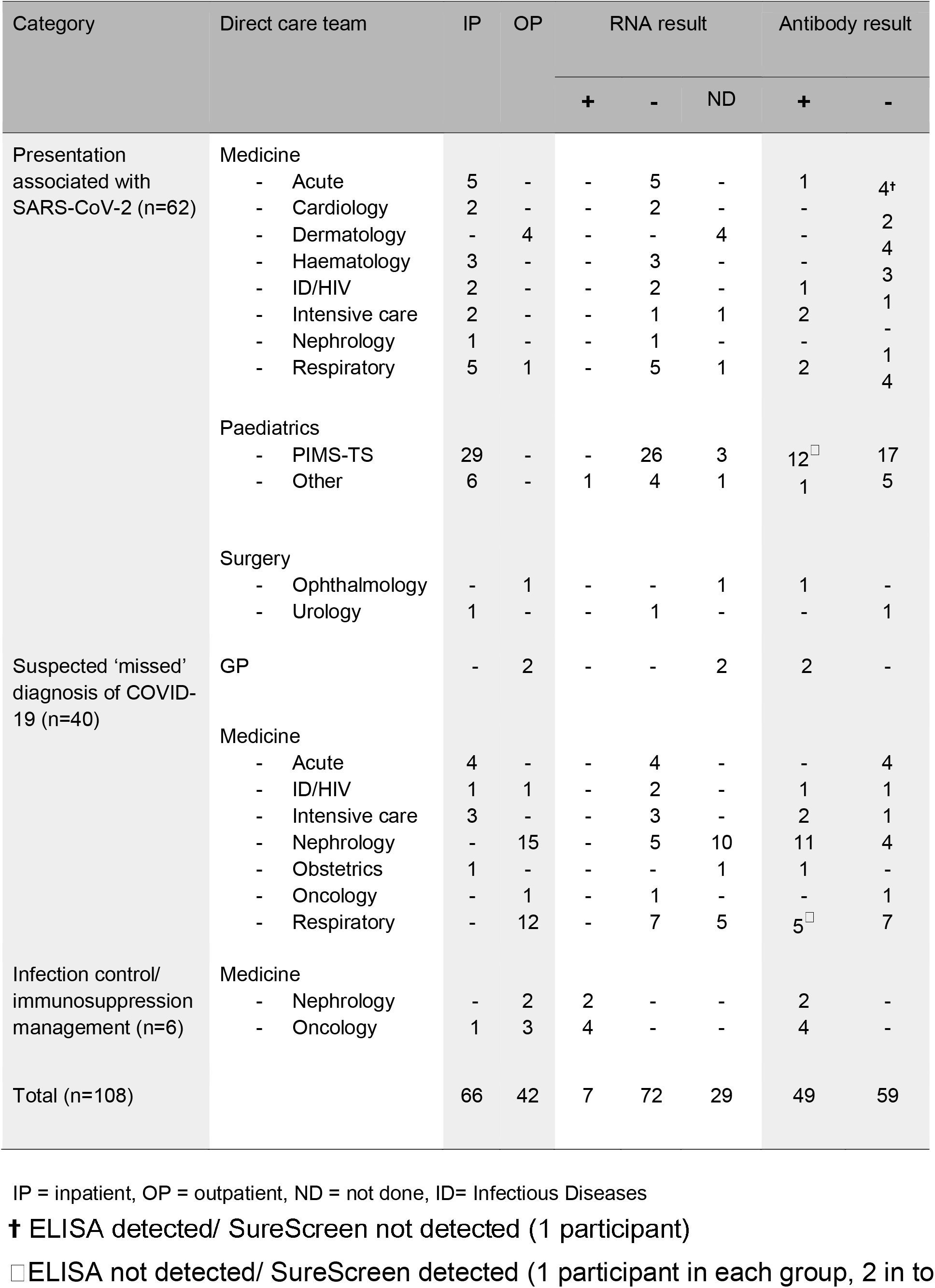
Referral characteristics and RNA results of individuals having SARS-CoV-2 serology testing performed during the pilot

Of 29 children with suspected PIMS-TS, 12 had detectable antibodies (41%). Reviewing the clinical history of the 17 with no detectable antibodies, seven (41%) had an alternate plausible diagnosis, or did not fulfill PIMS-TS diagnostic criteria at the time of discharge and 10 (59%) had ongoing high clinical suspicion of PIMS-TS. Two children had repeat testing, neither had detectable antibodies at this stage. For the remaining 33 RNA PCR negative individuals presenting with a potential post-COVID syndrome, seven (21.2%) had antibodies detected. This included two with the diagnosis of pulmonary embolism (PE), one with a new diagnosis of interstitial lung disease (ILD), two with a hyperinflammatory syndrome (akin to PIMS-TS), and one patient with paracentral acute middle maculopathy.

40 individuals were tested to identify potential missed COVID-19 diagnoses comprising nine presenting to hospital with ongoing compatible symptoms but negative SARS-CoV-2 RNA tests, and 31 who had recovered from a recent compatible illness in the community, including 15 individuals with end-stage renal failure, who had been advised to shield, and 12 patients attending the respiratory led post-COVID clinic due to failure to return to their baseline level of function. Serological testing was performed no earlier than 21 days post onset of symptoms (POS), up to approximately 90 days POS. Overall, 22/40 (55.0%) had detectable antibodies, including two patients admitted to ITU but with repeatedly negative RNA results on upper and lower respiratory sampling.

Of the 6 individuals with persistent SARS-CoV-2 RNA on nose and throat swabs tested to guide infection control or immunosuppression decisions, all had detectable antibodies on SureScreen LFIA, and when tested, moderate (n=1), high (n=1), or potent (n=4) neutralising antibodies titres. This implied, when considered with other factors such as time from first positive RNA test, and threshold cycle for RNA detection, that they were no longer infectious, and had a degree of protection from reinfection.

ELISA testing confirmed the LFIA result in all but three samples - in two cases the ELISA did not detect antibodies (whereas the SureScreen LFIA did), and in one case antibodies were detected (whereas the SureScreen LFIA detected none) (see table 2).

## DISCUSSION

This pilot SARS-CoV-2 serology service was introduced two months after the peak of acute UK COVID-19 admissions and provided results on 108 patients over a three-week period. It included a large number of children presenting with a new hyperinflammatory, Kawasaki-like syndrome, termed PIMS-TS(8), to the on-site Evelina London Children’s Hospital that provides tertiary referral and regional specialist services. 41% had antibodies detected, lower than previously reported(8,9), potentially due to increased awareness and broadening of clinical evaluation criteria, supported by a number of children having this diagnosis removed from discharge coding.

Serology was particularly helpful aiding diagnosis and management of what is an increasing range of assumed COVID-19 triggered conditions.(10-15) For example, antibodies were detected in two patients presenting with a PE that was therefore considered a provoked event, limiting the need for additional investigations and reducing the period of anticoagulation. Negative serology also helped discount COVID-19 as a potential trigger for newly presenting conditions, which included acquired haemophilia A and a range of unusual dermatological presentations e.g. ‘Covid toes’.

Detecting antibodies in patients with persistently positive SARS-CoV-2 PCR tests despite symptom resolution, a phenomenon reported elsewhere(16), enabled important decisions for infection control and immunosuppression. These decisions were supported by data that antibodies against spike protein (personal communication with SureScreen Diagnostics Ltd) correlate with neutralization(17) and there is published guidance that neutralisation can be used as a proxy for reduced risk of transmission (18,19). Since neutralising experiments are time-consuming and complex, rapid tests that detect antibodies against spike, such as the SureScreen LFIA and some, but not other technologies(20,21) are a practical alternative (22) when considered alongside other factors including timing from symptom onset, ongoing symptoms, and cycle threshold or take-off values of PCR results.

The strength of this study includes the extensive prior comparison of multiple technologies using a large panel of serum samples to inform choice and validation of the selected LFIA for clinical service. Results were also consistent with recommendations from a Cochrane review published after completion of our pilot, which suggested a benefit for serology to confirm a COVID-19 diagnosis in patients who did not have SARS-CoV-2 RNA testing performed, or who had a negative result despite an ongoing high index of clinical suspicion.(3)

It was also offered across the hospital to assess the broad potential clinical utility. There were also only 3 cases where concordance was not seen between the LFIA and ELISA, consistent with the sensitivity estimates from our validation. With high pre-test probability (e.g. 45%), the positive predictive value (PPV) is 99.2%, with an acceptable negative predictive value (NPV) of 96.9%. However, it is of note that if testing were to be extended to a population where prevalence is low (e.g. 5%) the PPV falls to <90%. This re-enforces the importance of providing serology for defined patient cohorts where the pre-test probability is high and the potential clinical utility is understood.

The main limitation of this study is in being performed at a single-centre at a discrete time-point in the COVID-19 pandemic. It is impossible to predict precise future serology service needs, whether that be aiding acute diagnosis alongside PCR testing,(23) informing patients that they have or have not had COVID-19, helping guide infection control decisions in hospitals or helping diagnose emerging post-inflammatory syndromes. Faster, more accurate even point of care SARS-CoV-2 virus detection assays may become widely available in hospitals and the community during a second wave, reducing the number of missed or delayed diagnoses and thus reduce demand for serological testing. The second limitation, although not technology specific, is that using serology as the marker of previous SARS-CoV-2 infection will likely only identify a proportion of infected patients given emerging evidence that seronegative individuals can show T-cell specific responses.(24)

Nevertheless, LFIAs are quick (10 minute test), straightforward to perform by trained operators, inexpensive and are already used in many diagnostic laboratories for example for detecting pneumococcal and legionella urinary antigens. They could also potentially be deployed outside pathology laboratories to provide more immediate results for decision making. This could include community healthcare facilities after appropriate training and mechanisms to record and disseminate the results. It should be noted however that as only serum was tested in our study, validation of LFIAs on capillary blood requires further work.

In summary, we conclude there is clinical utility in providing a SARS-CoV-2 patient serology service detecting spike proteins that can be feasibly delivered using LFIA devices. Further service evaluation at other centres will help track the emerging utility of serology testing and inform guidance on the indications and interpretation. There is a compelling case to provide such testing as soon as possible given emerging evidence(25) of a rapid decline in antibody levels, particularly in pauci-symptomatic patients.

## Data Availability

Deidentified participant data will be available, upon request, from corresponding author.

## Acknowledgements

We are extremely grateful to all staff in Viapath Infection Sciences and Department of Infectious Diseases based at St Thomas’ Hospital who helped deliver this service

## Contributors

NS and BM (equally) and RPG, SP, HW, AWS, GB SJDN, MGM, KD, EM, SD, RB GN and JDE conceived the study and study design. NS, BM, JR, EM, SD,GN, JDE screened and collated participant data. BM, RPG, SP, HW, AWS, GB, JR collected serum samples and conducted lateral flow experiments. KD designed ELISA and neutralising assay. KD, NK, SA,CG, JS conducted ELISA and neutralising assay experiments. NS and BM (equally) wrote the first draft of the manuscript. JDE is the guarantor. All authors interpreted the data and wrote and critically reviewed the manuscript and all revisions. The corresponding author attests that all listed authors meet authorship criteria and that no others meeting the criteria have been omitted.

## Ethical approva

All work was performed in accordance with the UK Policy Framework for Health and Social Care Research, and approved by the Risk and Assurance Committee at Guy’s and St Thomas’ NHS Foundation Trust. Informed consents were not required from participants in this study as per the guidelines set out in the UK Policy Framework for Health and Social Care Research and by the registration with, and express consent of the host institution’s review board.

## Data sharing

Data can be requested from the corresponding author.

## Guarantor Statement

The study guarantor (JDE) affirms that this manuscript is an honest, accurate, and transparent account of the study being reported; that no important aspects of the study have been omitted; and that any discrepancies from the study as planned (and, if relevant, registered) have been explained. Dissemination to participants and related patient and public communities: The results of the meta-analysis will be disseminated to patients, providers, policy makers through social media, and academic and institutional networks.

## Copyright/license for publication

This is an Open Access article distributed in accordance with the Creative Commons Attribution Non Commercial (CC BY-NC 4.0) license, which permits others to distribute, remix, adapt, build upon this work non-commercially, and license their derivative works on different terms, provided the original work is properly cited and the use is noncommercial. See: http://creativecommons.org/licenses/by-nc/4.0/

